# A gene expression map of host immune response in human brucellosis

**DOI:** 10.1101/2022.03.01.22271720

**Authors:** Ioannis Mitroulis, Akrivi Chrysanthopoulou, Georgios Divolis, Charalampos Ioannidis, Maria Ntinopoulou, Athanasios Tasis, Theocharis Konstantinidis, Natalia Soteriou, George Lallas, Stella Mitka, Mathias Lesche, Andreas Dahl, Stephanie Gembardt, Maria Panopoulou, Paschalis Sideras, Ben Wielockx, Ünal Coskun, Konstantinos Ritis, Panagiotis Skendros

## Abstract

Brucellosis is a common zoonotic disease caused by intracellular pathogens of the genus Brucella. Brucella infects macrophages and evades clearance mechanisms, which results in chronic parasitism. Herein, we studied the molecular changes that take place in human brucellosis both *in vitro* and *in vivo*. RNA sequencing was performed in primary human macrophages (Μφ) and polymorphonuclear neutrophils (PMNs) infected with clinical strains of *B. melitensis*. We observed a downregulation in the expression of genes involved in host response, such as TNF signaling, IL-1β production and phagosome formation in Μφ, and phosphatidylinositol signaling and TNF signaling in PMNs, being in line with the ability of the pathogen to survive within phagocytes. Further transcriptomic analysis of isolated peripheral blood mononuclear cells (PBMCs) and PMNs from patients with acute brucellosis before treatment initiation and after successful treatment revealed a positive correlation of the molecular signature of active disease with pathways associated with response to interferons (IFN). We identified 24 common genes that were significantly altered in both PMNs and PBMCs, including genes involved in IFN signaling that were downregulated after treatment in both cell populations, and *IL1R1* that was upregulated. The levels of several inflammatory mediators were evaluated in the serum of these patients, and we observed increased levels of IFN-γ, IL-1β and IL-6 before the treatment of acute brucellosis. An independent cohort of patients with chronic brucellosis also revealed the increased levels of IFN-γ during relapse compared to remissions. Taken together, this study provides for the first time an in-depth analysis of the molecular alterations that take place in human phagocytes upon infection, and in peripheral blood immune populations during active disease.

## INTRODUCTION

Brucellosis is a common bacterial zoonotic disease worldwide and an emerging zoonosis in several developed countries (1,2). Despite its importance in public health remains widespread and neglected in many areas, including southeastern Europe, Asia, Central and Latin America, and Africa (2,3). It is caused by intracellular pathogens of the genus Brucella that have domestic animals, mainly goats, sheep and cows, as natural reservoirs. The disease is transmitted to humans by consumption of unpasteurized milk and dairy products or by occupational contact with infected animals (4). Additionally, Brucella is highly infectious through the aerosol route, thus is considered as one of the most common laboratory-acquired pathogens and is also classified as a category B agent on the biodefense list (5,6).

Human brucellosis causes high morbidity and protean clinical manifestations, mimicking many infectious and non-infectious diseases since it can affect multiple organs. Despite early diagnosis and prolonged therapy with antibiotics is associated with substantial residual disability (4,7). Up to 30% of patients develop chronic disease, which is characterized by atypical clinical manifestations, high frequency of focal complications such as spondylitis, chronic fatigue syndrome, and relapses (7,8). Host protection against Brucella and prevention of its intracellular parasitism in macrophages depends on cell-mediated immunity, involving adequate Th1 immune response, with significant production of interferon gamma (IFN-γ) (8). Recent data support also an emerging key role of innate immunity and neutrophils in early proinflammatory responses against Brucella that may affect T-cell dynamics during infection (9,10). On the other hand, Brucella has developed various stealthy strategies to evade innate and adaptive immune responses, in order to establish intracellular long-term survival and replication (9,11). Several studies have demonstrated that patients with chronic brucellosis display defective cell-mediated immunity (brucellosis-acquired cellular anergy) probably due to modulation of host cellular immunity by Brucella (12). However, immunopathogenesis of human brucellosis remains incompletely understood and integrated molecular data that characterize complex interactions between Brucella and host immunity are missing today.

Here, we provide a gene expression map that guides the molecular signature of macrophages (Μφ) and polymorphonuclear neutrophils (PMNs) during the crucial early events of Brucella infection. Moreover, we analyse the transcriptomic alterations that take place concomitantly in peripheral blood mononuclear cells (PBMCs) and PMNs of patients upon treatment uncovering candidate molecular targets and pathways that may characterize active infection and disease eradication.

## MATERIALS AND METHODS

### Patients

Ten adult patients with acute brucellosis were recruited. EDTA anticoagulated blood and serum were collected from patients with active brucellosis before the initiation of antibiotic treatment and three months after the completion of treatment, when all patients were successfully treated. The diagnosis was based on compatible clinical manifestations in combination with high serum titers of anti-brucellar antibodies (Wright’s agglutination test ≥160) or a four-fold increase of the initial titers in two-paired samples drawn 2 weeks apart, or/and Brucella isolation. None of these patients suffered any relapse during a six-month post-treatment follow-up period. Patient characteristics and treatment are described in Table 1. PBMCs and PMNs were simultaneously isolated. RNA was isolated using TRIzol reagent (ThermoScientific). Sera from a second cohort of 25 chronic relapsing brucellosis patients at clinical relapse and remission, were also used. These patients had a disease duration of ≥12 months in combination with positive serum agglutination tests (SATs) or/and complement fixation test, or/and Brucella isolation (Supplementary Table 1).

**Table 1.**
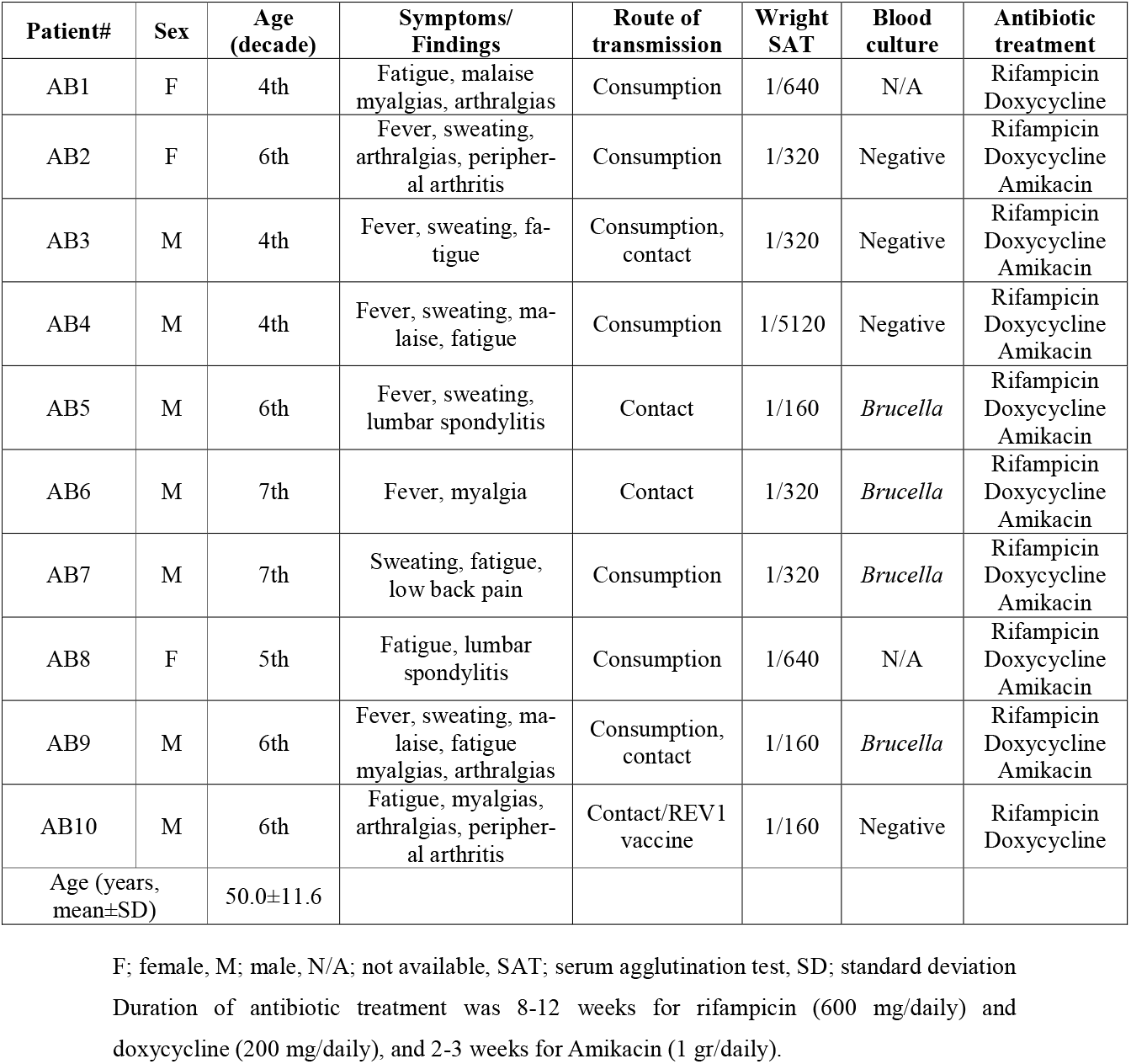
Characteristics of patients with acute brucellosis (AB).

Exclusion criteria were co-existence of other infectious, neoplasmatic or autoimmune disease, administration of immunomodulating agents or vaccination for at least 4 weeks before the entry to study, and pregnancy. The study was approved by the Local Scientific and Ethics Committee of the University Hospital of Alexandroupolis, Greece. All subjects provided written informed consent in accordance with the principles expressed in the Declaration of Helsinki.

### Mφ differentiation

Human Mφ were differentiated from isolated PBMCs from four control subjects, as previously described with modifications (13). In brief, PBMCs were isolated from healthy donors by density gradient centrifugation, using Histopaque 1077 (Sigma). To promote Mφ differentiation, monocytes were isolated using plastic adherence and cells were cultured in the presence of 10% autologous serum for 6 days.

### *In vitro* infection

Clinical strains of *B. melitensis* were used for *in vitro* experiments. Bacterial inoculum for cell infection was cultured on blood agar for 3 days. Bacterial suspension with 0.5 McFarland was opsonized for 30 minutes using human serum and then diluted in RPMI and ∼ 10^7^ bacteria in 0.5 ml of RPMI were added to each well (20 MOI) of PMNs or Mφ. Subsequently, cells were cultured for 0.5h for PMNs and 2h and 24h for Mφ, washed twice and RNA was isolated, using TRIzol reagent (ThermoScientific). Untreated PMNs and untreated Mφ, cultured for 2h served as control, respectively. The experimental procedure with *B. melitensis* was performed at biosafety level 3.

### RNA sequencing

RNA sequencing for Mφ and PBMCs was performed as previously described (14). To analyze RNA sequencing data, fragments were aligned with GSNAP (2020-12-16) to the human reference (hg38), and Ensembl annotation version 98 was used for the splice site support. Uniquely aligned fragments were counted with featureCounts (subread v2.0.1), again with the support of the Ensembl annotation. The exploratory analysis was performed with the DESeq 2 (v1.24.0) package within R (v3.6.3). Bias for patients was assessed using an exploratory correction with the variance stabilized transformation data of DESeq2 and the removeBatchEffect function of edgeR (3.26.8). Differential expression between before and after treatment was performed with a correction for patient.

For PMNs, 1000 ng of total RNA were used for the preparation of cDNA libraries, using the TruSeq RNA Library Preparation Kit v2 (Illumina), according to the manufacturer’s instructions. Library quality was evaluated using the Agilent DNA 1000 Kit (Agilent) with an Agilent 2100 Bioanalyzer. Quantification was performed by amplifying a set of six pre-diluted DNA standards (KAPA Biosystems) and diluted cDNA libraries by RT-qPCR. Isomolar quantities of up to 20 cDNA libraries, barcoded with different adaptors, were multiplexed. Sequencing was performed in a single-end manner at the Greek Genome Center, using a NextSeq 500/550 75c kit (Illumina) for the in vitro samples and a NovaSeq 6000 SP 100c kit (Illumina) for the ex vivo samples, generating 75 bp and 100 bp long reads, respectively, and an average of 25 million reads per library. Raw sequence data in FastQ format were uploaded to the Galaxy web platform, and standard tools of the public server “usegalaxy.org” were used for subsequent analysis (15). Briefly, quality control of raw reads was performed with FastQC (v072+galaxy1), followed by removal of adapter sequences and low-quality bases using Trim Galore! (v0.6.3). Next, HISAT2 (v2.2.1+galaxy0) was applied for the alignment of trimmed reads to the Homo sapiens (human) genome assembly GRCh37 (hg19) from Genome Reference Consortium. Assessment of uniform read coverage for exclusion of 5’/3’ bias and evaluation of RNA integrity at the transcript level were performed using Gene Body Coverage (v2.6.4.3) and Transcript Integrity Number (v2.6.4.1) tools, respectively. Differential gene expression was determined with DESeq2 (v2.11.40.6+galaxy1), using the count tables generated from HTSeq-count (v0.9.1) as input. The variability within and between individuals of this paired-data study was incorporated in the analysis, considering the treatment as the primary factor and the individual/patient as the secondary factor affecting gene expression.

Pathway and biological processes analysis was performed using the Enrichr analysis tool (14,16). Heat maps were generated using the Morpheus software (Broad Institute). Gene set enrichment (GSEA) pre-ranked analysis (1000 permutations, minimum term size of 15, maximum term size of 500) was performed using the GSEA software (Broad Institute). Gene sets were ranked by taking the -log10 transform of the p-value and signed as positive or negative based on the direction of fold change. Annotated gene sets from Molecular Signatures Database (MSigDB) were used as input (16).

### Cytokine measurement

The levels of cytokines were measured using the LEGENDplex™ Multi-Analyte Flow Assay Kit (Biolegend) in a CyFlow Cube 8 flow cytometer (Sysmex Partec, Germany), according to manufacturer’s instructions. For comparisons between the groups the Wilcoxon signed-rank test for paired samples was used. Statistical analysis was performed using GraphPad Prism (GraphPad Inc., La Jolla, CA). Significance was set at p < 0.05.

## RESULTS

### Analysis of the molecular signature of human macrophages infected *in vitro* with *B. melitensis*

To provide a time-course analysis of the molecular alterations of human Mφ during infection with *B. melitensis*, we performed *in vitro* infection of human Mφ, derived from the differentiation of peripheral blood monocytes from control subjects, and compared the transcriptomic signature of untreated Mφ compared to that of infected cells at 2h and 24h post-infection. Principal component analysis (PCA) revealed that there was a prominent change in the transcriptomic profile of Mφ at 24h after infection compared to untreated cells and cells at 2h after infection (Figure 1A). Pathway analysis, using the Kyoto Encyclopedia of Genes and Genomes (KEGG) database, of the significantly upregulated differentially expressed genes (DEG) (False Discovery Rate/FDR <0.01) between untreated Mφ and Mφ at 2h post-infection revealed an overrepresentation of circadian rhythm and ribosome biogenesis pathways, whereas downregulated DEGs were enriched in pathways associated with viral infection and infection from intracellular pathogens processes, including herpes simplex virus 1 infection, hepatitis C, and salmonella infection and TNF signaling (Figure 1B, C). Interestingly, we observed a decreased expression of genes encoding proteins critical in pathogen recognition, such as *NOD1, TLR5, TLR6* and *NLRC4* (Figure 1C).

**Figure 1.**
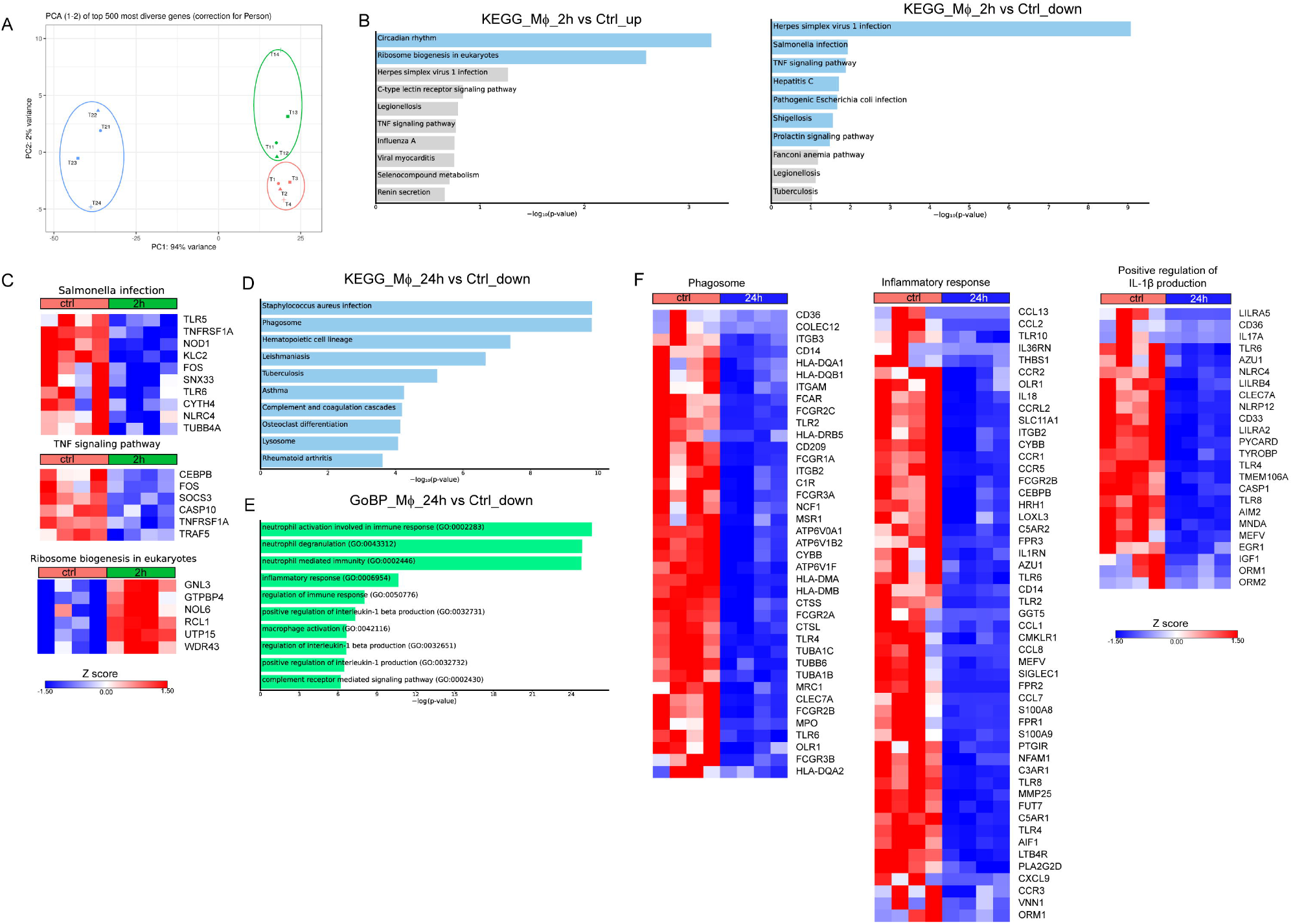
Alterations in the transcriptomic profile of human Mφ infected *in vitro* with *B. melitensis*. (A) Principal component analysis (PCA) of the transcriptome of all 12 Mφ samples. T1-T4 represent untreated control Mφ, T11-T14 represent samples from Mφ at 2h post infection and T21-T24 represent samples at the 24h time point. (B) Pathway analysis of the DEGs at 2h post-infection compared to control, using the KEGG database as reference. Light blue color represents statistical significance (C) Heatmaps depicting the DEGs of the respective pathways. (D) Pathway analysis of the DEGs with the highest variance at 24h post infection compared to control, using the KEGG database as reference. (E) Enriched biological processes in which the downregulated genes are involved. (F) Heatmaps depicting the DEGs of the phagosome pathway, the inflammatory response and positive regulation of IL-1β production biological processes.

We next assessed the molecular changes that take place at 24h post-infection. Pathway analysis of the downregulated DEGs with the highest variance (log2 fold change > 2 and < -2, FDR<0.01) showed overrepresentation of pathways associated with infection with Staphylococcus aureus and infection with intracellular pathogens, such as leishmaniasis and tuberculosis, as well as the pathways associated with phagosome and lysosome (Figure 1D). No statistically significant pathway was observed at the respective analysis of upregulated genes. Further, analysis of the DEGs that were downregulated at 24h after infection revealed that they are involved in biological processes associated with inflammation, and more specifically with the production of IL-1 and Mφ function (Figure 1E). Regarding the genes involved in the aforementioned pathways, there was a downregulation of several genes involved in the phagosome formation and function at 24h after infection, including those encoding for several Fcγ receptors (*FCGR1A, FCGR2A, FCGR2B, FCGR2C, FCGR3A, FCGR3B*), toll-like receptors (*TLR2, TLR4, TLR6*), other sensors of pathogen-associated molecular patterns (*CLEC7A, CD14*), integrins and other receptors involved in phagocytosis (*ITGB3, ITGAM, ITGB2, CD36*) (Figure 3D). We also observed a downregulation in the expression of genes encoding cytokines and cytokine receptors of the IL-1 family (*IL18, IL1RN, IL36RN*), chemokines (*CCL1, CCL2, CCL7, CCL8, CCL13, CXCL9*) and chemokine receptors (*CCR1, CCR2, CCR3, CCR5*), formyl peptide receptors (*FPR1, FPR2, FPR3*), and complement anaphylatoxin receptors (*C3AR1, C5AR1, C5AR2*) (Figure 1F). Regarding the regulation of IL-1 production, we observed the downregulation of several genes encoding inflammasome sensors (*NLRC4, NLRP12, MEFV, AIM2)*, the adaptor *PYCARD*, and the gene that encodes the effector *CASP1* (Figure 1F). Taken together, infection of Mφ with *B. melitensis* drives major changes in the transcriptomic profile of infected Mφ, downregulating a plethora of genes involved in the formation of phagosomes and the recognition of pathogens, in an effort to preserve pathogen survival within Mφ.

### Analysis of the molecular signature of human PMNs infected *in vitro* with *B. melitensis*

Even though Mφ are the major cell population infected by Brucella spp, it has been previously shown that this pathogen can also infect neutrophils (17). In order to characterize the molecular signature of infected PMNs with *B. melitensis*, we performed in vitro infection of human PMNs for 0.5h, derived from control subjects, and compared the transcriptomic signature of untreated PMNs to that of infected cells. Pathway analysis of the significantly overexpressed DEGs (FDR<0.01), using the KEGG database, highlighted Ribosome as the top upregulated pathway in Brucella-infected PMNs (Figure 2A). Notably, almost all genes (75 out of the 79) encoding for structural proteins of both small and large subunits of cytoplasmic ribosomes were found significantly upregulated (Figure 2B). Respective analysis of the downregulated DEGs demonstrated modulation of several pathways, some of which were also downregulated in Brucella-infected Μφ at 2h post-infection, such as TNF signaling and herpes simplex virus 1 infection (Figures 1B and 2C). However, various inflammation-related biological processes were significantly downregulated selectively in PMNs, namely the phosphatidylinositol signaling, NF-kappa B signaling, and cellular senescence pathways (Figure 2B, C). Amongst the downregulated transcripts in Brucella-infected PMNs, we identified several modulators of apoptosis (*BIRC3, FOXO3, DNM1L, ITPR1, TRAF1, TRAF5*) and inflammation, as exemplified by decreased mRNA expression of cytokines and corresponding receptors of the IL-1 family (*IL1A, IL1B, IL18R1*), chemokines (eg. *CCL20*), and various signaling mediators, such as kinases (*AKT3, ATM, ATR, CDK6, DGKD, DGKE, IPMK, IPPK, MAPK13, MAPK14, RPK1*) (Figure 2C).

**Figure 2.**
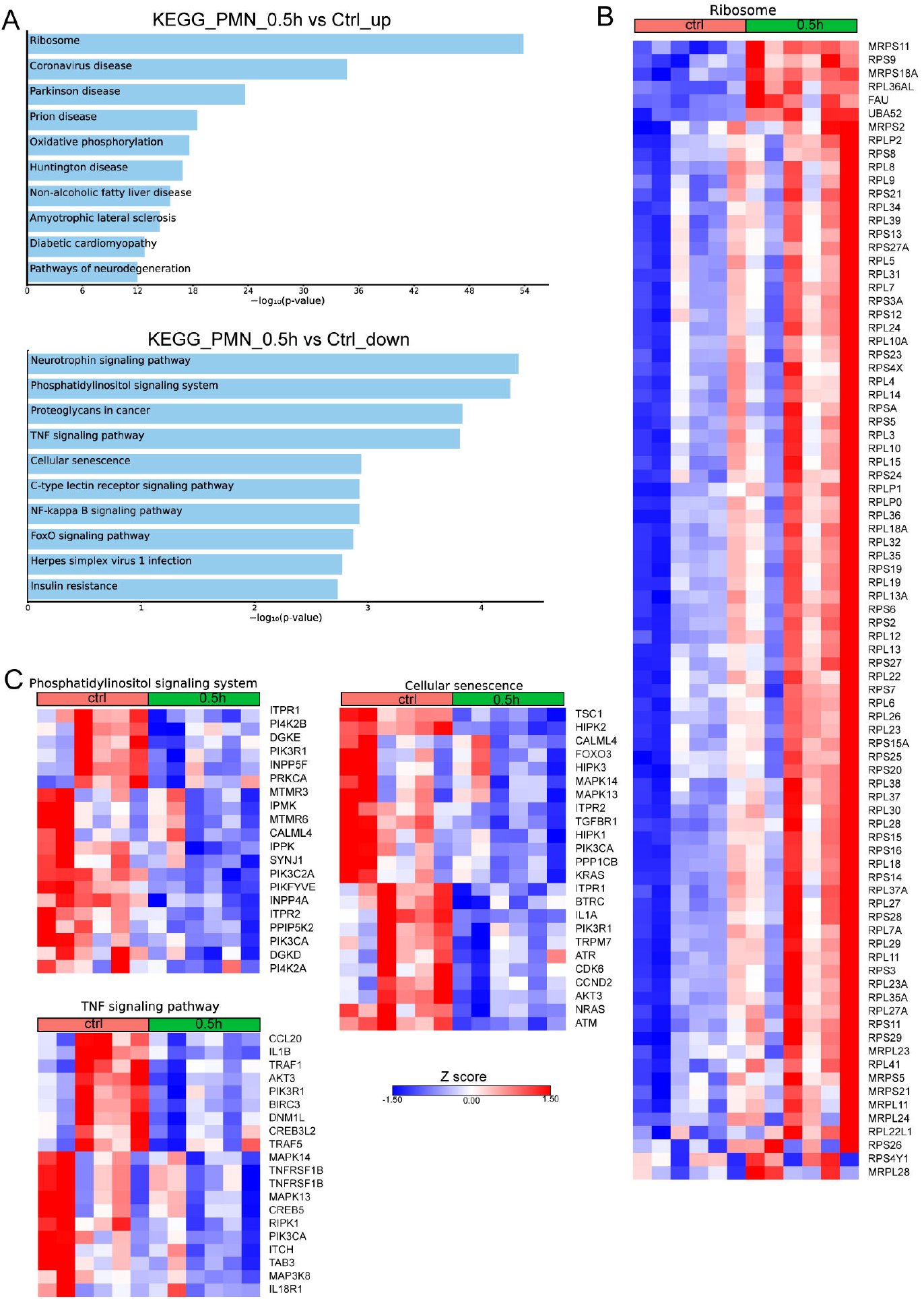
Alterations in the transcriptomic profile of human PMNs infected in vitro with *B. melitensis*. (A) Pathway analysis of the DEGs from PMNs at 0.5h post infection with *B. melitensis* compared to control, using the KEGG database as reference. (B) Heatmap depicting the DEGs of the ribosome pathway. (C) Heatmap depicting the DEGs of the pathways enriched for downregulated genes.

### Transcriptomic profiling of active human brucellosis

We further investigated the transcriptomic signature of active human brucellosis. To do so, PMNs were isolated from eight patients with active brucellosis before the initiation of antibiotic treatment (active disease) and three months after completion of the antibiotic treatment, when patients were free of symptoms (remission). Transcriptomic analysis identified 318 DEGs (FDR<0.1) (Figure 3A). DEGs that were upregulated after treatment are involved in RNA transport and autophagy pathways, whereas downregulated DEGs after treatment are involved in NOD-like receptor signaling pathway and cytokine-cytokine receptor interaction pathways, as well as several pathways associated with infectious diseases (Figure 3A). The upregulated genes that encode proteins involved in RNA transport were the members of the eukaryotic initiator factors (EIF) family *EIF1, EIF3I, EIF4A3, EIF5*, and the genes of the autophagy pathway were *ATG2A, GABARAPL1, TP53INP2, DDIT4* and *IRS2* (Figure 3B). On the other hand, we observed a downregulation of critical genes in immune regulation, such as *IL1B, CX3CR1, CCR2, CCR5, CXCR6, STAT1, AIM2*, and *CD40*, as well as genes associated with interferon signaling, such as *OAS1, OAS2, GBP1* and *GPB3* (Figure 3B). We further performed gene set enrichment analysis (GSEA) using the Hallmark Gene Set collection of the Molecular Signatures Database. We observed a positive correlation of the transcriptomic signature of PMNs during active brucellosis with IFN-γ and IFN-α response and with inflammatory response (Figure 3).

**Figure 3.**
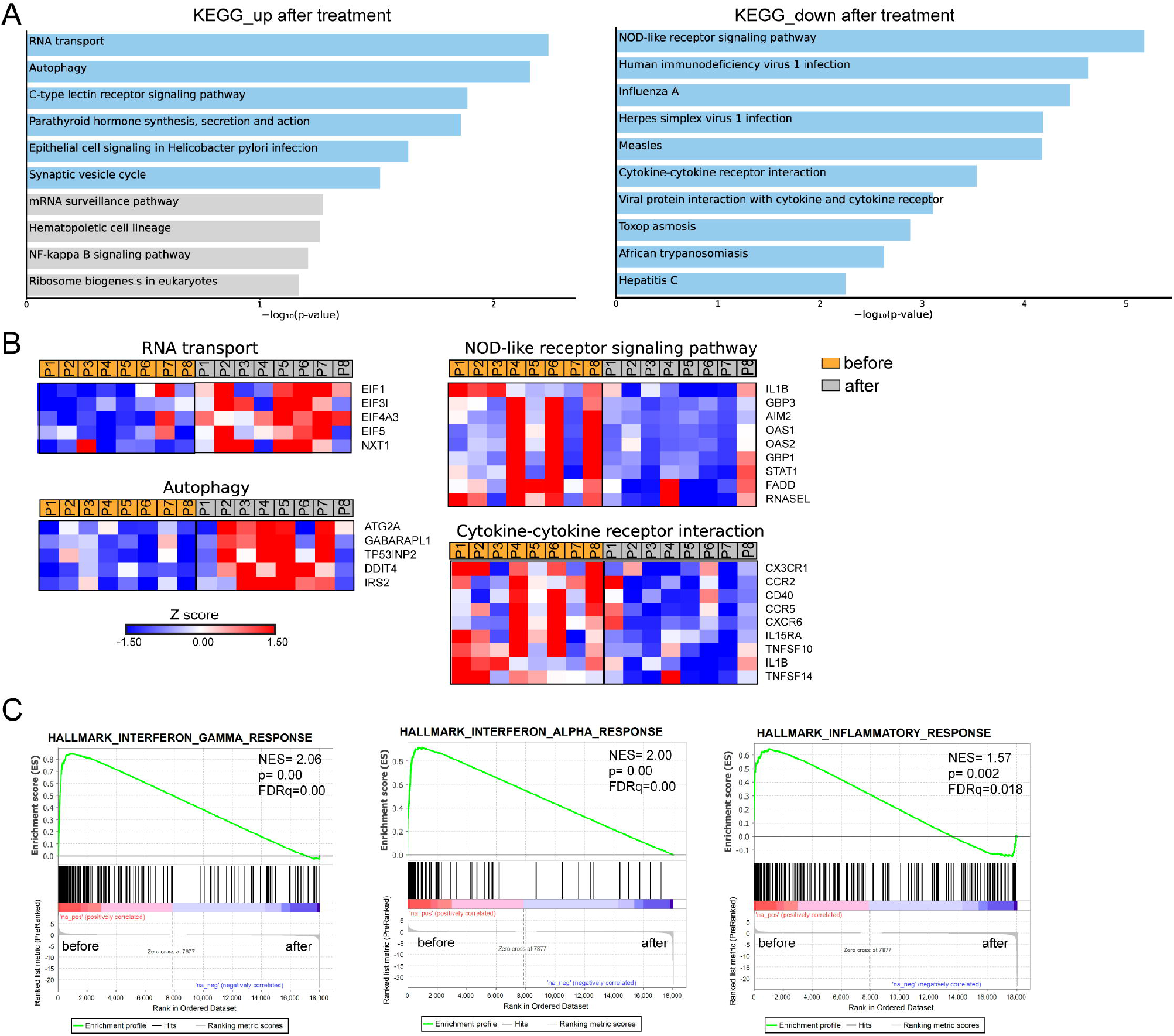
Transcriptomic analysis of PMNs from patients with brucellosis before treatment initiation and after successful completion of treatment. (A) Pathway analysis of the DEGs from PMNs after treatment compared to PMNs isolated from the same patients (paired-data analysis) during active brucellosis, using the KEGG database as reference. Light blue color represents statistical significance (B) Heatmaps depicting the DEGs of the respective pathways. P1-P8 refers to different patients. (C) GSEA for genes related to response to interferons, and inflammation.

In parallel, we performed transcriptomic analysis of PBMCs isolated from six patients with active brucellosis before and after antibiotic treatment. Transcriptomic analysis identified 62 genes with significantly altered expression (FDR<0.1) after treatment (Figure 4A). We observed that successful treatment resulted in the increased expression of *HIF1A*, a critical regulator of inflammation, and of the genes that encode IL-1 receptor *IL1R1*, and its accessory protein *IL1RAP*, which form a complex that mediates IL-1 signal transduction (Figure 4A). On the other hand, there was a downregulation in the expression of genes that play a major role in immune function, such as *CD274*, which encodes PD-L1, *STAT1, CD3G*, the intracellular immunoglobulin receptor *TRIM21, CXCR6*, the lymphocytic activation molecules *SLMF6, SLAMF7* and genes that encode proteins important in effector cell cytolytic processes, such as *CD160, GZMA, GZMH* (Figure 4A). Moreover, several identified genes are involved in interferon-related activation pathways, such as *GPB3, GPB4, OAS1, OAS2, OASL, IFI16*, and *XAF1* (Figure 4A). To the same line, GSEA analysis revealed that the gene sets with the most significant positive association with active disease were IFN signaling and OXPHOS, whereas the one with the most significant negative association was the hypoxia gene set (Figure 4B). Notably, we further identified 24 genes that were differentially expressed both in PMNs and PBMCs (Figure 4C). Among these common genes, *CXCR6, TRIM21, SLAM7, CD274* and the genes associated with IFN signaling *OASL, OAS1, OAS2, GBP3*, and *STAT1* were downregulated in both datasets, whereas *IL1R1* was commonly upregulated (Figure 4D).

**Figure 4.**
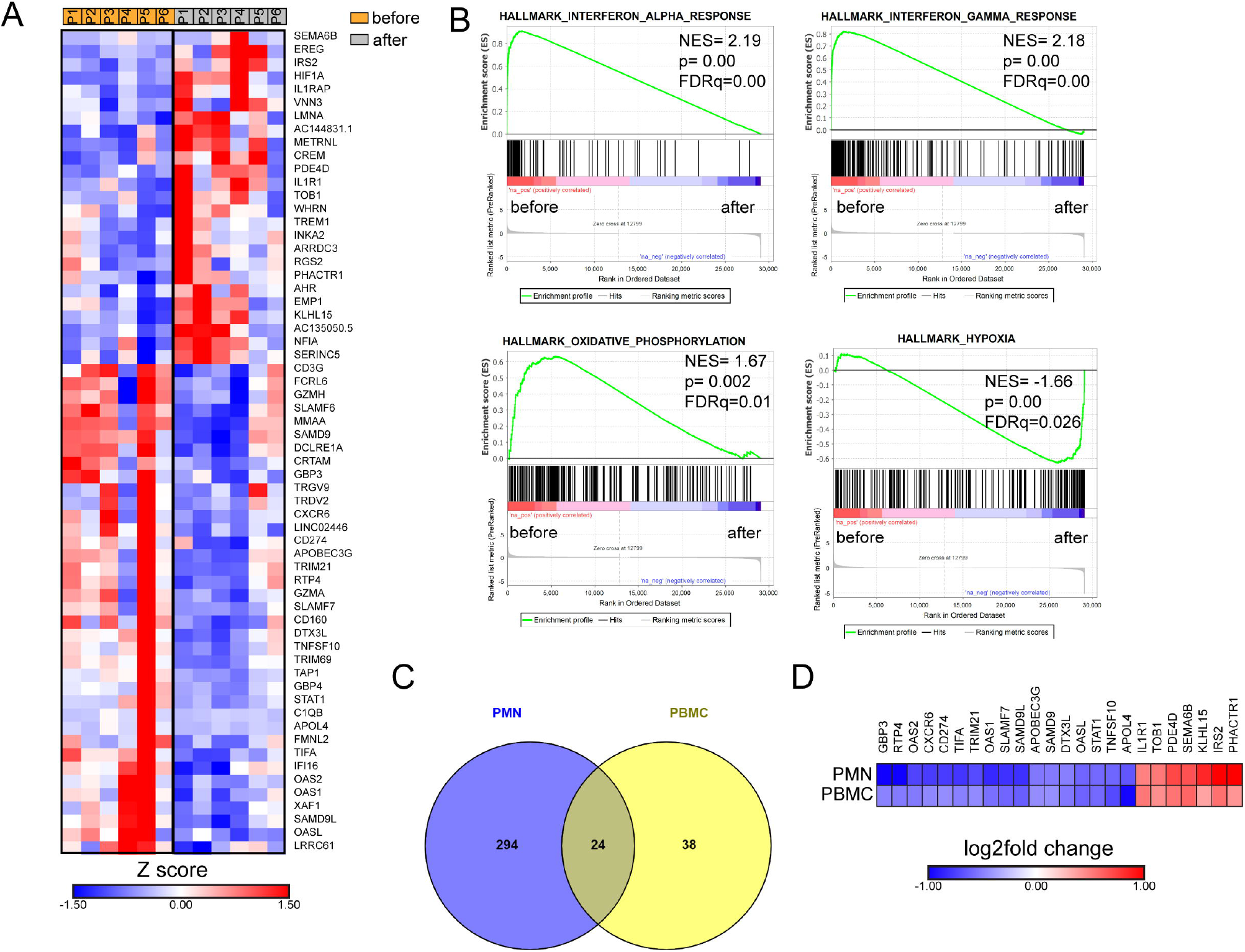
Transcriptomic analysis of PBMCs from patients with brucellosis before treatment initiation and after successful completion of treatment. (A) Heatmap depicting the DEGs from PBMCs from patients with acute brucellosis before treatment initiation and from the same patients (paired analysis) after successful treatment. P1-P6 refers to different patients. (B) GSEA for genes related to response to interferons, oxidative phosphorylation and hypoxia. (C) Venn diagram and (D) heatmap depicting the common genes that were significantly differentially expressed in PMNs and PBMCs from patients with brucellosis after treatment.

### Cytokine levels in acute brucellosis

To this point, we observed that the molecular signature that characterizes acute brucellosis is positively correlated with those of IFN-α and IFN-γ response. For this reason, we measured the levels of several cytokines in the sera of patients during acute brucellosis and after successful treatment. We observed a significant downregulation in the levels of IFN-γ, IL-1β and IL-6 post-treatment, whereas there was no statistically significant difference in the levels of IFN-α, IL-18, TNF, MCP-1 and IL-17A (Figure 5A-H). We further confirmed that the levels of IFN-γ are increased in active disease in a cohort of patients with chronic relapsing brucellosis. In this cohort, the levels of IFN-γ were increased during relapse compared to remission (Figure 5I).

**Figure 5.**
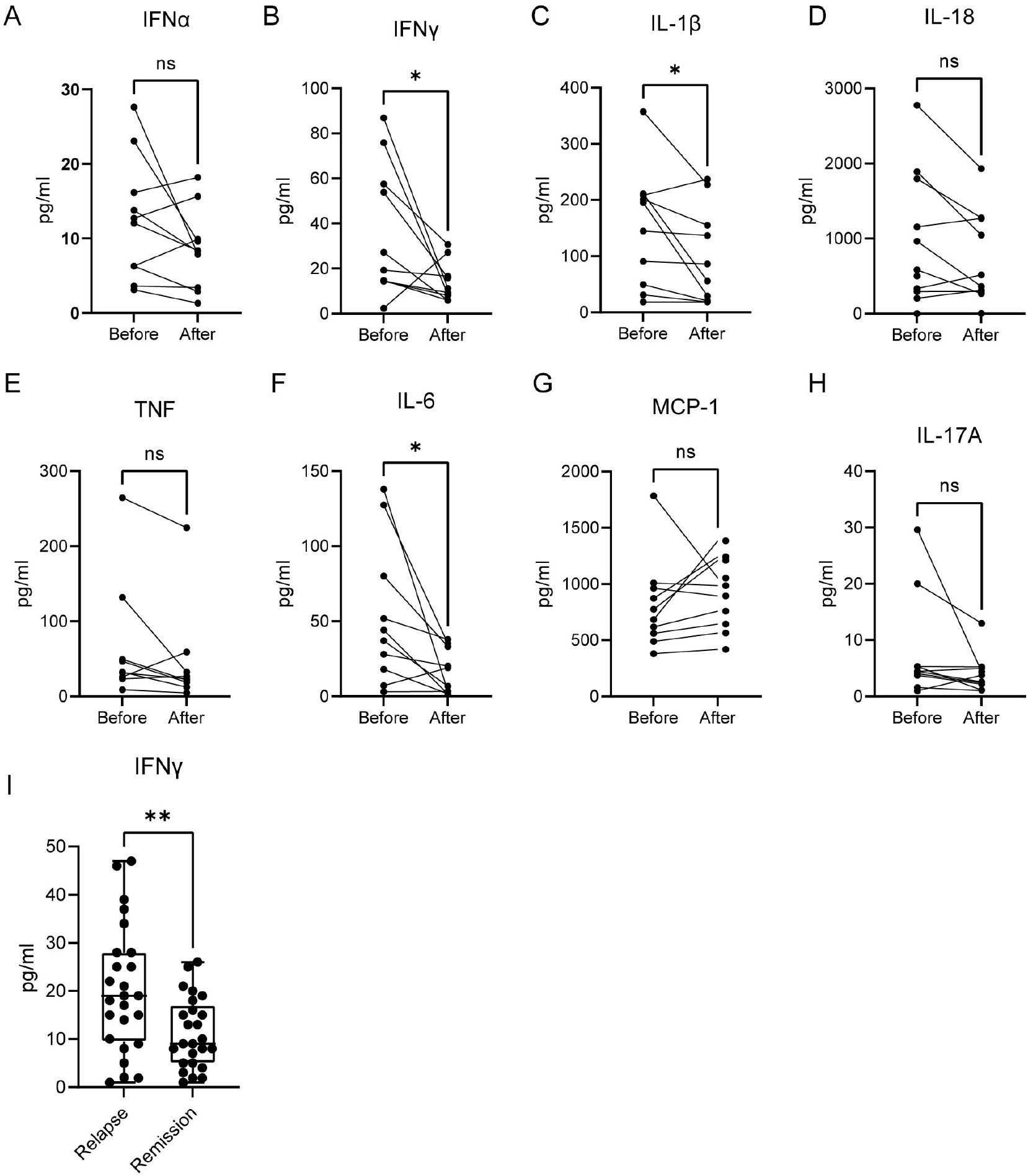
Levels of cytokines in the serum of patients with active brucellosis. (A-H) Levels of IFN-α, IFN-γ, IL-1β, IL-18, TNF, IL-6, MCP-1 and IL-17A in the serum of patients with acute brucellosis before treatment initiation and after successful treatment. (I) Levels of IFN-γ in patients with chronic relapsing brucellosis during relapse and remission. *p<0.05, **p<0.01. Wilcoxon signed rank test.

## DISCUSSION

The interaction between Brucella and host immune system is critical for the development of persistent infection or infection clearance (8,18). To date, transcriptomic data derived from Brucella-infected mouse macrophages or mouse cell lines, domestic ruminants or Brucella-vaccinated animals (19–25). This study analyses, for the first time, the transcriptome profile, both *in vitro*, in Brucella-infected primary Mφ and PMNs, and *ex-vivo*, in PBMCs and PMNs derived from patients with acute brucellosis before and after treatment. This provides the molecular signature that characterizes the main host cellular immune populations during their initial interplay with invading Brucella, and the molecular signature of different stages of the disease.

Early molecular events following phagocytosis of Brucella by macrophages are crucial for the activation of innate immunity leading to the induction of a favorable Th1 response (8,18). Several lines of evidence indicated that Brucella manipulates multiple effector mechanisms in macrophages to its own benefit (9,12). In line with this, we identified that in Mφ infected *in vitro* by a clinical strain of *B. melitensis*, the expression of several genes encoding key proteins involved in the recognition of Brucella and in the proinflammatory response against the pathogen were markedly suppressed. These alterations may initiate as soon as 2h post-infection being more prominent at 24h post infection. Interestingly, most downregulated DEGs related to phagosome, TNFa signaling and IL-1β production. Indeed, previous studies reported that various Brucella virulence factors and pathogen-associated molecular patterns (PAMPs), such as Type IV secretory system (T4SS), lipopolysaccharide (LPS) and outer membrane lipoproteins (OMPs) modify phagosome biogenesis and trafficking in macrophages to inhibit phagolysosome fusion, and develop suitable vacuolar compartments to enable intracellular replication of the microbe (9,12). Moreover, the current study comes in agreement with previous data demonstrated that Brucella Omp25 protein inhibits *in vitro* the production of TNF in human Μφ and dendritic cells preventing cell maturation and antigen presentation (26–28). Furthermore, several genes encoding members of the IL-1 family (*IL18, IL1RN, IL36RN*) and inflammasome complexes (*NLRC4, NLRP12, MEFV, AIM2, PYCARD, CASP1)* are significantly downregulated in Brucella-infected Μφ. Experimental studies indicated that inflammasomes and their effectors are essential for an initial effective immune response against Brucella infection (29–31). On the other hand, Brucella can regulate canonical and non-canonical inflammasome signaling and pyroptosis in macrophages by impairing caspase-1 and caspase-4/11 activation, and IL-1β secretion (32,33). It is intriguing that Brucella downregulates macrophage *MEFV* expression, the gene responsible for familial Mediterranean fever, the prototype IL-1β-mediated autoinflammatory disease (34). Mutations in the *MEFV* gene are highly prevalent in the Middle East and Mediterranean countries where brucellosis is endemic (35). Our data further support the hypothesis that *MEFV* mutations may provide an evolutionary selective advantage to confer protection against brucellosis (36).

Recently, PMNs emerge as novel players during the initial stages of innate immune response against Brucella infection (10). Brucella resists the killing mechanisms of human PMNs and induces the early death of these cells promoting their phagocytosis by Mφ, which become vehicles for bacterial dispersion within the host (37). Studies in murine brucellosis proposed that infected PMNs attenuate cellular adaptive immunity, given that depletion of PMNs favored bacterial elimination (17,38). Based on these, this study examined the early transcriptome alterations of *in vitro* Brucella-infected neutrophils, before their premature death. *B. melitensis*-infected PMNs were characterized by increased expression of genes associated with ribosome biogenesis indicating bulk induction of protein synthesis, in an effort to arm their bactericidal mechanisms and survive. Of interest and in a similar way to Mφ, *in vitro* infection of PMNs with Brucella led to downregulated gene expression in key molecular pathways for PMNs physiology and function including phosphatidylinositol signaling, TNF signaling, and cellular senescence. Phosphatidylinositol signaling pathway plays an important role in membrane dynamics and trafficking, including proteins implicated in endosomal membranes and autophagosome assembly and activity (39,40). Autophagy is closely related to the intracellular lifestyle of many pathogens, including Brucella (41,42). We hypothesize that the downregulation of several autophagy sensors and regulators belonging to phosphatidylinositol pathway further modulates the autophagic capacity of PMNs against Brucella. This may also explain the inability of Brucella-infected PMNs to form neutrophil extracellular traps (NETs) (17), an effector mechanism positively associated with the autophagy machinery (43). Downregulation of the cellular senescent pathway is in agreement with the reported premature death of Brucella-infected PMNs (17). Additionally, senescence has been associated with resistance to cellular death (44). Moreover, it appears that perturbation of TNF signaling represents a common stealth strategy of Brucella to avoid both Mφ- and PMNs-induced inflammation further restricting cellular immunity (11).

In order to investigate the impact of human brucellosis in host immunity and identify possible candidate markers of active disease and response to treatment, we next assessed the transcriptome profiling of PBMCs and PMNs isolated from newly diagnosed patients with acute brucellosis, before and three months after their successful treatment. We observed, both in PBMCs and PMNs, transcriptomic alterations related to major pathways of inflammation, supporting its role for infection overcome. PBMCs from patients successfully treated were characterized by the overexpression of genes critically involved in hypoxia (*HIF1A*) and IL-1 signaling, and the downregulation of genes implicated in oxidative phosphorylation, lymphocyte activation, and cytotoxicity. In line with these data, a recent experimental study has demonstrated that absence of HIF-1α renders mice susceptible to Brucella infection, while HIF-1α reduces oxidative phosphorylation and increases glycolysis leading to inflammasome activation and IL-1β release in infected macrophages (45).

Treatment of brucellosis upregulated in PMNs the expression of several genes related to autophagy machinery, including *DDIT4/REDD1* encoding a key regulator of autophagy-mediated NET formation (47). It seems that after clearance of infection, PMNs restored critical functions impaired by Brucella, such as autophagy. However, they did not acquire a proinflammatory phenotype as indicated by the downregulated expression in genes related to pathways that contain NOD-like receptor signaling and cytokine-cytokine interactions.

Of note, this study identified a common set of 24 genes that were differentially expressed both in PMNs and PBMCs suggesting candidate molecular diagnostic/prognostic targets for human brucellosis. Among them, IFN pathway, which is the major driver of Th1 immunity against Brucella (8), appears to be induced in active disease and attenuated after treatment. Indeed, using patients’ sera, we confirm at the protein level, that IFN-γ and other Th1 cytokines, such as IL-1β and IL-6, were increased during active disease and significantly diminished in cured, non-relapsed patients. Collectively, these results confirmed past studies highlighting the significant role of a robust Th1 response to tackle acute infection and brucellosis-acquired cellular anergy of chronic disease (47–50).

In conclusion, this study provides an integrated transcriptome landscape of immune cells signature in human brucellosis suggesting candidate molecular pathways and targets for active disease and response to treatment. Based on these data, future validation and mechanistic studies may further decipher the pathogenesis of this ancient and continuously re-emerging zoonotic disease (1,2,51).

## Data Availability

All data produced in the present study are available upon reasonable request to the authors

## Acknowledgements

This study was supported by the German Federal Ministry for Education and Research (BMBF) and Greek General Secretariat for Research and Technology (GSRT), German-Greek Bilateral Research & Innovation Programme BRIDGING, grant MIS 5030062, and partially was supported by the GSRT, Research & Innovation Programme CYTONET, grant MIS 5048548.

## References

1. Dean AS, Crump L, Greter H, Schelling E, Zinsstag J. Global burden of human brucellosis: a systematic review of disease frequency. PLoS Negl Trop Dis. 2012;6(10):e1865.

2. European Food Safety Authority and European Centre for Disease Prevention and Control (EFSA and ECDC). The European Union summary report on trends and sources of zoonoses, zoonotic agents and food-borne outbreaks in 2017. EFSA J. 2018 Dec;16(12):e05500.

3. Franc KA, Krecek RC, Häsler BN, Arenas-Gamboa AM. Brucellosis remains a neglected disease in the developing world: a call for interdisciplinary action. BMC Public Health. 2018 Jan 11;18(1):125.

4. Pappas G, Akritidis N, Bosilkovski M, Tsianos E. Brucellosis. N Engl J Med. 2005 Jun 2;352(22):2325–36.

5. Pereira CR, Cotrim de Almeida JVF, Cardoso de Oliveira IR, Faria de Oliveira L, Pereira LJ, Zangerônimo MG, et al. Occupational exposure to Brucella spp.: A systematic review and meta-analysis. PLoS Negl Trop Dis. 2020 May;14(5):e0008164.

6. Pappas G, Panagopoulou P, Christou L, Akritidis N. Brucella as a biological weapon. Cell Mol Life Sci. 2006 Oct;63(19–20):2229–36.

7. Dean AS, Crump L, Greter H, Hattendorf J, Schelling E, Zinsstag J. Clinical manifestations of human brucellosis: a systematic review and meta-analysis. PLoS Negl Trop Dis. 2012;6(12):e1929.

8. Skendros P, Pappas G, Boura P. Cell-mediated immunity in human brucellosis. Microbes Infect. 2011 Feb;13(2):134–42.

9. Jiao H, Zhou Z, Li B, Xiao Y, Li M, Zeng H, et al. The Mechanism of Facultative Intracellular Parasitism of Brucella. Int J Mol Sci. 2021 Apr 1;22(7):3673.

10. Moreno E, Barquero-Calvo E. The Role of Neutrophils in Brucellosis. Microbiol Mol Biol Rev. 2020 Nov 18;84(4):e00048–20.

11. Martirosyan A, Moreno E, Gorvel J-P. An evolutionary strategy for a stealthy intracellular Brucella pathogen. Immunol Rev. 2011 Mar;240(1):211–34.

12. Skendros P, Boura P. Immunity to brucellosis. Rev Sci Tech. 2013 Apr;32(1):137–47.

13. Kourtzelis I, Li X, Mitroulis I, Grosser D, Kajikawa T, Wang B, et al. DEL-1 promotes macrophage efferocytosis and clearance of inflammation. Nat Immunol. 2019 Jan;20(1):40–9.

14. Mitroulis I, Ruppova K, Wang B, Chen L-S, Grzybek M, Grinenko T, et al. Modulation of Myelopoiesis Progenitors Is an Integral Component of Trained Immunity. Cell. 2018 Jan 11;172(1–2):147–161.e12.

15. Afgan E, Baker D, Batut B, van den Beek M, Bouvier D, Cech M, et al. The Galaxy platform for accessible, reproducible and collaborative biomedical analyses: 2018 update. Nucleic Acids Res. 2018 Jul 2;46(W1):W537–44.

16. Lamprianidou E, Kordella C, Kazachenka A, Zoulia E, Bernard E, Filia A, et al. Modulation of IL-6/STAT3 signaling axis in CD4+FOXP3-T cells represents a potential antitumor mechanism of azacitidine. Blood Adv. 2021 Jan 12;5(1):129–42.

17. Barquero-Calvo E, Martirosyan A, Ordoñez-Rueda D, Arce-Gorvel V, Alfaro-Alarcón A, Lepidi H, et al. Neutrophils exert a suppressive effect on Th1 responses to intracellular pathogen Brucella abortus. PLoS Pathog. 2013 Feb;9(2):e1003167.

18. de Figueiredo P, Ficht TA, Rice-Ficht A, Rossetti CA, Adams LG. Pathogenesis and immunobiology of brucellosis: review of Brucella-host interactions. Am J Pathol. 2015 Jun;185(6):1505–17.

19. Zhou Z, Gu G, Luo Y, Li W, Li B, Zhao Y, et al. Immunological pathways of macrophage response to Brucella ovis infection. Innate Immun. 2020 Oct;26(7):635–48.

20. Zhou D, Zhi F, Fang J, Zheng W, Li J, Zhang G, et al. RNA-Seq Analysis Reveals the Role of Omp16 in Brucella-Infected RAW264.7 Cells. Front Vet Sci. 2021;8:646839.

21. Solanki KS, Gandham RK, Thomas P, Chaudhuri P. Transcriptome analysis of Brucella abortus S19Δper immunized mouse spleen revealed activation of MHC-I and MHC-II pathways. Access Microbiol. 2020;2(1):acmi000082.

22. Hop HT, Arayan LT, Reyes AWB, Huy TXN, Min W, Lee HJ, et al. Simultaneous RNA-seq based transcriptional profiling of intracellular Brucella abortus and B. abortus-infected murine macrophages. Microb Pathog. 2017 Dec;113:57–67.

23. Liu Q, Han W, Sun C, Zhou L, Ma L, Lei L, et al. Deep sequencing-based expression transcriptional profiling changes during Brucella infection. Microb Pathog. 2012 Dec;52(5):267–77.

24. Rossetti CA, Galindo CL, Everts RE, Lewin HA, Garner HR, Adams LG. Comparative analysis of the early transcriptome of Brucella abortus--infected monocyte-derived macrophages from cattle naturally resistant or susceptible to brucellosis. Res Vet Sci. 2011 Aug;91(1):40–51.

25. Wang F, Hu S, Liu W, Qiao Z, Gao Y, Bu Z. Deep-sequencing analysis of the mouse transcriptome response to infection with Brucella melitensis strains of differing virulence. PLoS One. 2011;6(12):e28485.

26. Jubier-Maurin V, Boigegrain RA, Cloeckaert A, Gross A, Alvarez-Martinez MT, Terraza A, et al. Major outer membrane protein Omp25 of Brucella suis is involved in inhibition of tumor necrosis factor alpha production during infection of human macrophages. Infect Immun. 2001 Aug;69(8):4823–30.

27. Caron E, Gross A, Liautard JP, Dornand J. Brucella species release a specific, protease-sensitive, inhibitor of TNF-alpha expression, active on human macrophage-like cells. J Immunol. 1996 Apr 15;156(8):2885–93.

28. Billard E, Dornand J, Gross A. Brucella suis prevents human dendritic cell maturation and antigen presentation through regulation of tumor necrosis factor alpha secretion. Infect Immun. 2007 Oct;75(10):4980–9.

29. Costa Franco MMS, Marim FM, Alves-Silva J, Cerqueira D, Rungue M, Tavares IP, et al. AIM2 senses Brucella abortus DNA in dendritic cells to induce IL-1β secretion, pyroptosis and resistance to bacterial infection in mice. Microbes Infect. 2019 Mar;21(2):85–93.

30. Lacey CA, Mitchell WJ, Dadelahi AS, Skyberg JA. Caspase-1 and Caspase-11 Mediate Pyroptosis, Inflammation, and Control of Brucella Joint Infection. Infect Immun. 2018 Sep;86(9):e00361–18.

31. Gomes MTR, Campos PC, Oliveira FS, Corsetti PP, Bortoluci KR, Cunha LD, et al. Critical role of ASC inflammasomes and bacterial type IV secretion system in caspase-1 activation and host innate resistance to Brucella abortus infection. J Immunol. 2013 Apr 1;190(7):3629–38.

32. Jakka P, Namani S, Murugan S, Rai N, Radhakrishnan G. The Brucella effector protein TcpB induces degradation of inflammatory caspases and thereby subverts non-canonical inflammasome activation in macrophages. J Biol Chem. 2017 Dec 15;292(50):20613–27.

33. Campos PC, Gomes MTR, Marinho FAV, Guimarães ES, de Moura Lodi Cruz MGF, Oliveira SC. Brucella abortus nitric oxide metabolite regulates inflammasome activation and IL-1β secretion in murine macrophages. Eur J Immunol. 2019 Jul;49(7):1023–37.

34. Skendros P, Papagoras C, Mitroulis I, Ritis K. Autoinflammation: Lessons from the study of familial Mediterranean fever. J Autoimmun. 2019 Nov;104:102305.

35. Özen S. Update on the epidemiology and disease outcome of Familial Mediterranean fever. Best Pract Res Clin Rheumatol. 2018 Apr;32(2):254–60.

36. Ross JJ. Goats, germs, and fever: Are the pyrin mutations responsible for familial Mediterranean fever protective against Brucellosis? Med Hypotheses. 2007;68(3):499–501.

37. Gutiérrez-Jiménez C, Mora-Cartín R, Altamirano-Silva P, Chacón-Díaz C, Chaves-Olarte E, Moreno E, et al. Neutrophils as Trojan Horse Vehicles for Brucella abortus Macrophage Infection. Front Immunol. 2019;10:1012.

38. Mora-Cartín R, Gutiérrez-Jiménez C, Alfaro-Alarcón A, Chaves-Olarte E, Chacón-Díaz C, Barquero-Calvo E, et al. Neutrophils Dampen Adaptive Immunity in Brucellosis. Infect Immun. 2019 Mar;87(5):e00118–19.

39. Baba T, Balla T. Emerging roles of phosphatidylinositol 4-phosphate and phosphatidylinositol 4,5-bisphosphate as regulators of multiple steps in autophagy. J Biochem. 2020 Oct 1;168(4):329–36.

40. Claude-Taupin A, Morel E. Phosphoinositides: Functions in autophagy-related stress responses. Biochim Biophys Acta Mol Cell Biol Lipids. 2021 Jun;1866(6):158903.

41. Starr T, Child R, Wehrly TD, Hansen B, Hwang S, López-Otin C, et al. Selective subversion of autophagy complexes facilitates completion of the Brucella intracellular cycle. Cell Host Microbe. 2012 Jan 19;11(1):33–45.

42. Skendros P, Mitroulis I. Host cell autophagy in immune response to zoonotic infections. Clin Dev Immunol. 2012;2012:910525.

43. Skendros P, Mitroulis I, Ritis K. Autophagy in Neutrophils: From Granulopoiesis to Neutrophil Extracellular Traps. Front Cell Dev Biol. 2018;6:109.

44. Sasaki M, Kumazaki T, Takano H, Nishiyama M, Mitsui Y. Senescent cells are resistant to death despite low Bcl-2 level. Mech Ageing Dev. 2001 Oct;122(15):1695–706.

45. Gomes MTR, Guimarães ES, Marinho FV, Macedo I, Aguiar ERGR, Barber GN, et al. STING regulates metabolic reprogramming in macrophages via HIF-1α during Brucella infection. PLoS Pathog. 2021 May;17(5):e1009597.

46. Skendros P, Chrysanthopoulou A, Rousset F, Kambas K, Arampatzioglou A, Mitsios A, et al. Regulated in development and DNA damage responses 1 (REDD1) links stress with IL-1β-mediated familial Mediterranean fever attack through autophagy-driven neutrophil extracellular traps. J Allergy Clin Immunol. 2017 Nov;140(5):1378–1387.e13.

47. Giambartolomei GH, Delpino MV, Cahanovich ME, Wallach JC, Baldi PC, Velikovsky CA, et al. Diminished production of T helper 1 cytokines correlates with T cell unresponsiveness to Brucella cytoplasmic proteins in chronic human brucellosis. J Infect Dis. 2002 Jul 15;186(2):252–9.

48. Rafiei A, Ardestani SK, Kariminia A, Keyhani A, Mohraz M, Amirkhani A. Dominant Th1 cytokine production in early onset of human brucellosis followed by switching towards Th2 along prolongation of disease. J Infect. 2006 Nov;53(5):315–24.

49. Skendros P, Boura P, Chrisagis D, Raptopoulou-Gigi M. Diminished percentage of CD4+ T-lymphocytes expressing interleukine-2 receptor alpha in chronic brucellosis. J Infect. 2007 Feb;54(2):192–7.

50. Skendros P, Sarantopoulos A, Tselios K, Boura P. Chronic brucellosis patients retain low frequency of CD4+ T-lymphocytes expressing CD25 and CD28 after Escherichia coli LPS stimulation of PHA-cultured PBMCs. Clin Dev Immunol. 2008;2008:327346.

51. Godfroid J, Cloeckaert A, Liautard J-P, Kohler S, Fretin D, Walravens K, et al. From the discovery of the Malta fever’s agent to the discovery of a marine mammal reservoir, brucellosis has continuously been a re-emerging zoonosis. Vet Res. 2005 Jun;36(3):313–26.

