## Supplementary Table 1 for "A gene expression map of host immune response in human brucellosis"

### SUPPLEMENTARY MATERIAL

**Supplementary Table 1.** Diagnostic laboratory findings at relapse and remission of patients with chronic relapsing brucellosis (24 males/1 female, mean age 60.6±6.9 years).

| #Patient | Blood culture | | Wright's SAT (positive ≥1:160) | | Coombs' antibrucella SAT (positive ≥1:320) | | Complement Fixation test (positive ≥1:16) | |
|---|---|---|---|---|---|---|---|---|
| | **Rel** | **Rem** | **Rel** | **Rem** | **Rel** | **Rem** | **Rel** | **Rem** |
| **CRB1** | pos | neg | 160 | 160 | 640 | 640 | 32 | 32 |
| **CRB2** | neg | neg | 320 | 160 | >1280 | 640 | 32 | 16 |
| **CRB3** | pos | neg | 160 | 80 | 640 | 320 | 64 | 16 |
| **CRB4** | pos | neg | 80 | 80 | 320 | 320 | 64 | 32 |
| **CRB5** | neg | neg | 320 | 320 | >1280 | >1280 | 256 | 64 |
| **CRB6** | neg | neg | 160 | 160 | 640 | 640 | 128 | 64 |
| **CRB7** | pos | neg | 160 | 160 | 1280 | 320 | 128 | 16 |
| **CRB8** | neg | neg | 320 | 640 | >1280 | >1280 | 32 | 16 |
| **CRB9** | pos | neg | 160 | 160 | 640 | 640 | 128 | 32 |
| **CRB10** | neg | neg | 160 | 80 | 160 | 160 | 64 | 16 |
| **CRB11** | neg | neg | 80 | 160 | 1280 | 640 | 128 | 32 |
| **CRB12** | neg | neg | 160 | 80 | 320 | 320 | 16 | 32 |
| **CRB13** | pos | neg | 80 | 40 | 320 | 160 | 256 | 32 |
| **CRB14** | neg | neg | 80 | 160 | 320 | 640 | 128 | 64 |
| **CRB15** | neg | neg | 640 | 160 | >1280 | 640 | 64 | 64 |
| **CRB16** | neg | neg | 320 | 160 | 320 | 160 | 128 | 64 |
| **CRB18** | neg | neg | 320 | 640 | >1280 | >1280 | 32 | 16 |
| **CRB19** | pos | neg | 160 | 160 | 640 | 640 | 128 | 32 |
| **CRB20** | neg | neg | 160 | 80 | 160 | 160 | 64 | 16 |
| **CRB21** | neg | neg | 80 | 160 | 1280 | 640 | 128 | 32 |
| **CRB22** | neg | neg | 160 | 80 | 320 | 320 | 16 | 32 |
| **CRB23** | neg | neg | 80 | 160 | 320 | 640 | 128 | 64 |
| **CRB24** | neg | neg | 640 | 160 | >1280 | 640 | 64 | 64 |
| **CRB25** | pos | neg | 640 | 320 | 1280 | >1280 | >256 | 32 |

CRB; chronic relapsing brucellosis, rel; relapse, rem; remission, pos; positive, neg; negative

SAT; serum agglutination test. Results of serological tests are expressed as reciprocal titers.
